# Ultrasound-guided bleomycin sclerotherapy for Warthin’s tumor: preliminary results of a prospective study

**DOI:** 10.1101/2023.08.30.23294824

**Authors:** Eeva Castren, Minna Sirviö, Katri Aro, Kimmo Lappalainen, Riikka Lindén, Goran Kurdo, Jussi Tarkkanen, Antti Mäkitie, Timo Atula

**Author notes:** Correspondence: Eeva Castren.

## Abstract

**Background:** Warthin’s tumor (WT) is the second most common parotid gland neoplasm with rising incidence. As some WT patients are not suitable for surgery, non-surgical treatment options are needed. This is the first study to evaluate the feasibility of ultrasound-guided bleomycin sclerotherapy (UGBS) for WT.

**Patients and methods:** WT patients with concordant clinical, imaging, and cytology data were included. UGBS was proposed for patients who desired non-surgical intervention and fulfilled the inclusion criteria. Tumor volume and tumor-related symptoms were registered before UGBS and six months after.

**Results:** Nine patients underwent UGBS between September 2021 and September 2022. In seven of the nine patients (78%), the tumor volume had diminished on average by 34% at six-month follow-up. Also, tumor-related pain, and cosmetic and functional discomfort decreased during the follow-up. One patient was later referred to partial parotidectomy.

**Conclusion:** UGBS could be an alternative non-surgical treatment for WT, especially for patients with risk factors for surgery.

## Introduction

Warthin’s tumor (WT) is the second most common benign tumor of the parotid gland and typically occurs in older men with history of smoking. (1) Recent studies show a rising incidence of WT. (2-5) WT may be bilateral, the patients may have multiple lesions in the same gland and postoperative recurrences are encountered. (6,7)

Traditionally, the first-line treatment for WT has been superficial or partial parotidectomy. However, recent studies propose a more conservative approach, as the diagnosis of WT is accurate when based on concordant clinical, imaging and cytology data, and as its malignant transformation is considered extremely rare. (1,8,9) Less invasive treatment strategies are needed because many WT patients have relevant risk factors related to surgery: either elevated risk for general anesthesia due to comorbidities, or previous parotidectomy increasing the risk of facial nerve injury.

In this study, we prospectively assess whether ultrasound-guided bleomycin sclerotherapy (UGBS) could serve as a non-surgical treatment option for WT. It induces inflammation that gradually results in shrinkage of the lesion. UGBS has long been applied for low-flow vascular malformations locally, and it has been shown to be efficient with few reported complications. (10,11)

Currently, reports on non-surgical interventions for WT are limited. (12-15) This is the first study that prospectively evaluates the efficacy and safety of UGBS in the management of WTs.

## Patients and methods

We enrolled all patients diagnosed with WT at the Department of Otorhinolaryngology – Head and Neck Surgery (Helsinki, Finland) during September 2021 to September 2022. The diagnosis of WT was confirmed by typical clinical history, clinical behavior of the tumor, ultrasound (US) examination and fine or core needle biopsy assessed by a Head and Neck pathologist at our institution. Once the diagnosis of WT was confirmed, we informed the patients about treatment options: either intervention or active surveillance (AS), if the tumor was asymptomatic. The first line intervention was surgery but UGBS was proposed as an option in particular for patients who desired an intervention for the tumor, but declined surgery or had relevant risk factors for partial parotidectomy, i. e. previous surgery of the same gland increasing the risk of facial nerve injury, previous removal of another major salivary gland, or relevant risk for general anesthesia or other major surgical complications. Whether the patient was enrolled for surgery, UGBS, or AS was decided by the patient and the clinician.

For patients included in the study, we recorded demographic data, relevant medical history, history of smoking, previous WT, and salivary gland surgery. Tumor volume, location, morphology and cytology were recorded. At enrollment, the patients reported symptoms related to WT from 0 to 5 (0= asymptomatic, 1=very mild discomfort, 2=mild discomfort, 3=moderate discomfort, 4=relevant discomfort, 5=very relevant discomfort) regarding pain, cosmetic or functional discomfort (i.e. effect on eating, sleeping, shaving or other daily activities), or any other symptoms caused by the tumor.

The inclusion criteria for UGBS were: parotid gland neoplasm diagnosed as WT based on concordant clinical, imaging and cytology data, glomerular filtration rate (GFR) >50 ml/min and ability to undergo magnetic resonance imaging (MRI). We excluded WT patients who preferred surgery or AS, had fibrotic lung disease, acute pneumonia or substantially compromised lung function, GFR <50 ml/min, active cancer disease, allergy to bleomycin, or who were pregnant, breastfeeding or planning reproductive cell donation within six months after UGBS.

Patients enrolled for UGBS underwent MRI to further confirm the diagnosis of WT, as well as the size and morphology of the tumor, and to rule out other conditions in the parotid gland. GFR was measured at the time of enrollment and one week prior to UGBS.

### Ultrasound-guided bleomycin sclerotherapy

An experienced interventional radiologist familiar with UGBS performed the treatment. Tumor volume was measured, the puncture site was locally anesthetized and the tumor was first aspirated to drain its cystic component. Then, 15 000 IU bleomycin diluted in suitable volume of saline was injected into the tumor. In case of multiple tumors, the largest tumor was treated, except in one patient, who received the 15 000 IU of bleomycin divided into two equally sized tumors. The patients stayed under supervision for 30 minutes and were then discharged if no acute adverse events occurred.

To record any adverse events or complications after UGBS, the patients were advised to contact our institution in case of unexpected symptoms after UGBS. The patient records were controlled two weeks after the treatment and at the six-month follow-up for any complications.

### Follow-up

US was performed six months after UGBS to evaluate the size and morphology of the tumor. The patients were contacted to report tumor-related symptoms regarding cosmetic and functional discomfort, pain, and patient-reported alteration of tumor size. They were also asked whether they would recommend UGBS, and whether they desired further treatment at this stage of the follow-up. A one-year follow-up was planned for all patients.

### Ethical considerations

The study protocol was approved by the institutional Research Ethics Committee and by the Finnish Medical Agency (Fimea), and the protocol was registered in European Union Clinical Trials Register by EudraCT number 2021-001972-41. Enrolled patients were informed about the study design and they signed an informed consent.

### Statistical analysis

Statistical analysis was performed using Microsoft Excel and SPSS. Mann-Whitney U-test was applied to analyze statistical significance of tumor volume alteration and tumor-related symptoms before and after UGBS.

## Results

Between September 2021 and September 2022, 51 WT patients were enrolled in the study. Out of these 51 patients, nine opted for UGBS and fulfilled the inclusion criteria. By April 2023, these nine patients had completed their six-month follow-up. Most patients were men (n=7) and aged on average 67 years (range 57-80). They all had a history of smoking and eight out of the nine were current smokers. All had histories of cardiovascular disease, diabetes mellitus, chronic obstructive pulmonary disease, or sleep apnea. Bilateral WT was observed in two patients and multiple tumors were observed in five. The reason for opting for UGBS instead of surgery was the patient’s fear of complications related to surgery or a relevant risk for general anesthesia. None had had previous parotidectomy.

### Tumor volume and tumor-related symptoms before and after UGBS

Table 1 shows in detail the tumor volumes and tumor-related symptoms before and six months after UGBS.

**Table 1.**
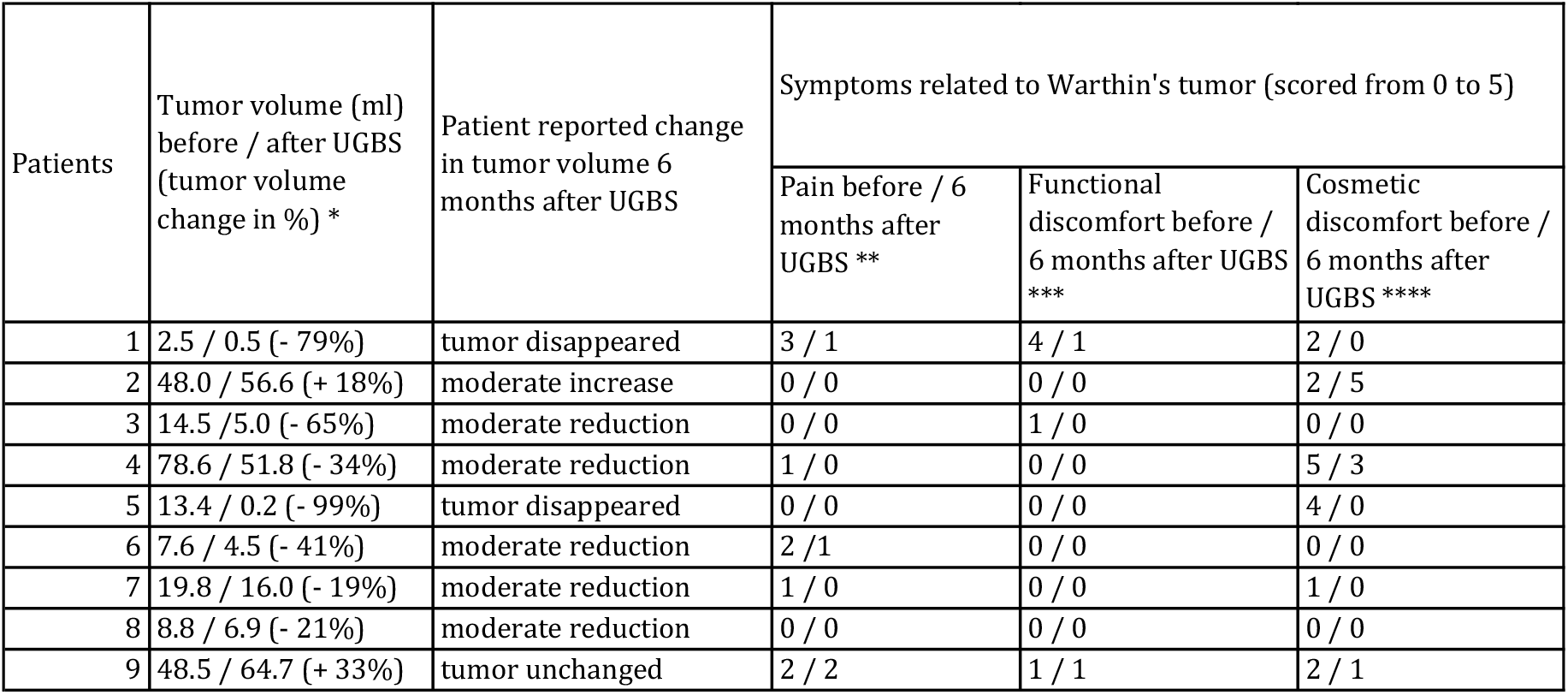
Summary of tumor volumes and symptoms related to WT before and 6 months after UGBS. Symptoms were scored from 0 to 5, (0= asymptomatic, 1=very mild discomfort, 2=mild discomfort, 3=moderate discomfort, 4=relevant discomfort, 5=very relevant discomfort * p=0.23 ** p=0.33 *** p=0.66 **** p=0.29

| Patients | Tumor volume (ml) before / after UGBS (tumor volume change in %) * | Patient reported change in tumor volume 6 months after UGBS | Symptoms related to Warthin's tumor (scored from 0 to 5) |  |  |
| --- | --- | --- | --- | --- | --- |
|  |  |  | Pain before / 6 months after UGBS ** | Functional discomfort before / 6 months after UGBS *** | Cosmetic discomfort before / 6 months after UGBS **** |
| 1 | 2.5 / 0.5 (- 79%) | tumor disappeared | 3 / 1 | 4 / 1 | 2 / 0 |
| 2 | 48.0 / 56.6 (+ 18%) | moderate increase | 0 / 0 | 0 / 0 | 2 / 5 |
| 3 | 14.5 / 5.0 (- 65%) | moderate reduction | 0 / 0 | 1 / 0 | 0 / 0 |
| 4 | 78.6 / 51.8 (- 34%) | moderate reduction | 1 / 0 | 0 / 0 | 5 / 3 |
| 5 | 13.4 / 0.2 (- 99%) | tumor disappeared | 0 / 0 | 0 / 0 | 4 / 0 |
| 6 | 7.6 / 4.5 (- 41%) | moderate reduction | 2 / 1 | 0 / 0 | 0 / 0 |
| 7 | 19.8 / 16.0 (- 19%) | moderate reduction | 1 / 0 | 0 / 0 | 1 / 0 |
| 8 | 8.8 / 6.9 (- 21%) | moderate reduction | 0 / 0 | 0 / 0 | 0 / 0 |
| 9 | 48.5 / 64.7 (+ 33%) | tumor unchanged | 2 / 2 | 1 / 1 | 2 / 1 |

Prior to intervention, the tumor volume, when calculated based on the largest tumor, was on average 26.9 ml, and tumor volumes ranged from 2.5 ml to 78.6 ml. At the six-month follow-up, tumor volume assessed by US decreased on average by 34.2%. In seven of the nine patients, the tumor volume diminished by 19-99%, and in two patients it increased by 18-33%. The alterations in tumor volume were not statistically significant. When asked about changes in tumor size at the six-month follow-up, most patients reported that the tumor had either disappeared or decreased moderately.

In eight of the nine of patients, tumor-related symptoms (scored from 0 to 5) decreased: cosmetic discomfort decreased on average from 1.8 to 1.0, functional discomfort from 0.7 to 0.2 and pain fell from 1.0 to 0.4. The alteration of symptoms was not statistically significant. Finally, eight of the nine patients would recommend this procedure as an alternative treatment for WT.

### Complications and further treatments

No acute adverse events occurred. One patient developed a bacterial infection of the puncture site three days after UGBS and needed drainage with needle and intravenous antibiotics (Clavien-Dindo complications grade II). The infection resolved but tumor volume increased in the follow-up, so he later underwent partial parotidectomy. One patient reported transient headache one week after UGBS. No skin or nerve complications or systemic adverse events occurred. At the six-month follow-up, 89% of the patients were satisfied and did not desire further surgery. One patient with bilateral WT is scheduled for a contralateral side UGBS.

## Discussion

This study is to our knowledge the first prospective study to evaluate the feasibility of ultrasound-guided bleomycin sclerotherapy as a non-surgical treatment option for WT. Our preliminary results show that in most WT patients, UGBS reduces the tumor volume, and most patients report less tumor-related symptoms after UGBS.

Traditionally, surgery has been the golden standard for parotid gland neoplasms, as cytology and imaging have been considered insufficient to rule out malignancy. However, current literature supports a more conservative approach: when clinical, state-of-the art imaging, and cytology data concordantly support the diagnosis of WT, it is considered very reliable, and surgery is not mandatory just to rule out malignancy. (8,9). Malignant transformation of WT has also been found to be extremely rare. (1) Therefore, it seems to be safe not to operate on WT, and surveillance or non-surgical treatments can be offered. (9) UGBS could be a treatment option for patients who do not want surgery, or bear a high risk for surgical complication, but report tumor-related discomfort.

To date, reports on non-surgical interventions on WT have been limited. However, the need of non-surgical treatments is obvious, as the incidence of WT is rising and some WT patients are unsuitable for surgery. Two case reports on ultrasound-guided ethanol sclerotherapy exist with positive results. (12,13) Ultrasound-guided microwave ablation (MWA) and radiofrequency ablation (RFA) have also been utilized in small sets of patient with promising results. (14,15) We opted for bleomycin, as it has been widely used with good results for low-flow vascular malformations with minimal complications and minimal nerve damage. (10,11) Bleomycin, compared to other sclerosants such as ethanol, has shown the least post-procedural oedema, skin complications, and neuropathy, which is crucial in proximity of the facial nerve. (10,11,16). The feared side-effect of bleomycin in oncological use is pulmonary fibrosis; however, bleomycin-induced pulmonary fibrosis has only been reported with cumulative doses higher than 150 000 IU in conjunction with compromised renal function and the simultaneous use of other cytostatics. (10,16) To our knowledge, pulmonary fibrosis has not been reported after UGBS for vascular malformations.

There are a few advantages in treating a WT with UGBS. First, UGBS is performed as an outpatient procedure under local anesthesia, and for many WT patients avoiding general anesthesia is relevant. As UGBS does not require an operating room or a hospital stay, the strain on the patient and the health care system is moderate. Second, patients with multiple or bilateral WTs could benefit from UGBS, especially if they have had previous salivary gland surgery: healthy salivary gland tissue can be spared and the risk of facial nerve injury through repeated surgery can be avoided. Third, based on our experience, previous UGBS does not seem to hinder future parotid gland surgery: one patient in this study and another patient outside of this study underwent partial parotidectomy after UGBS, and the course of surgery was uneventful.

Is UGBS efficient and safe for WT? To evaluate, whether UGBS is clinically efficient or not in the long run obviously calls for larger studies and longer follow-ups. However, the patient’s opinion on the result is crucial in addition to the US-assessed tumor volume change. Our results show that UGBS decreases tumor volume in most patients, not only as evaluated by US, but also as evaluated by the patient. Twenty-two percent of the patients reported that their tumors had disappeared, and 56% reported moderate tumor size reduction. We also observed reductions in tumor-related symptoms. However, these findings were not statistically significant, as the number of patients was limited. The follow-up period in this series is currently six months, but we will continue to follow whether the effect of UGBS is permanent. Previous case reports on ultrasound-guided ethanol sclerotherapy showed similar results in two patients: the tumor volume shrank by 55-98%, but follow-up time was only 2-5 months. (12,13) Sclerotherapy is considered less painful than RFA or MWA, which use thermal energy, and potential thermal damage to the facial nerve should also be considered. (12-15) One post-procedural infection occurred in the present series, which most likely was more due to the puncture itself. To date, we have not observed other relevant complications.

The results of UGBS are promising, but a larger study population and a longer follow-up period will eventually show the efficacy of UGBS. We will continue to follow-up the present series and enroll more patients to further assess the feasibility of UGBS for WT.

## Data Availability

All data produced in the present study are available upon reasonable request to the authors

